# Sialic Acid Receptors in Human Mammary Tissue: Implications for Infection with Novel Influenza Strains

**DOI:** 10.1101/2025.08.06.25333154

**Authors:** Carrie L. Byington, Donald Pizzo, Hana Russo, Steven L. Gonias

**Author notes:** Corresponding Author Contact Information: Carrie L. Byington, MD, Department of Pediatrics, University of California, San Diego 9461 Gilman Dr, La Jolla, CA 92093.

## Abstract

**Importance:** Highly pathogenic avian influenza (HPAI) A H5N1 has been recognized for nearly three decades as a threat to avian species and as a virus with pandemic potential if spillover into human populations occurs. Recently the virus has evolved capacity to infect many mammalian species, including dairy cattle, increasing the risk for human exposure and the pandemic threat. Sialic acids (SA) serve as binding sites for influenza viruses. The distribution of SA determines infectivity of specific influenza viruses across species and tissue tropism. Hemagglutinin (HA) of human and swine adapted influenza viruses bind primarily to SA with α2,6-galactose linkages and avian influenza viruses preferentially bind to SA with α 2,3-galactose linkages. Recently, the bovine udder was found to contain SA with α2,3 linkages which allow the H5N1 virus to bind to bovine udder epithelium and to infect milk. The distribution of SA receptors in the human mammary gland is unknown.

**Objective:** Evaluate normal human mammary tissue for the presence of both human and avian SA receptors.

**Design:** Retrospective evaluation of lectin binding to sialic acids in human mammary tissue. Setting: US academic health center.

**Participants:** Specimens obtained from de-identified women with residual non-malignant tissue following breast surgery at the University of California, San Diego.

**Exposures:** Lectin affinity-staining of human tissue for the presence of SA.

Main Outcomes and Measures: Presence or absence of lectin staining.

**Results:** All mammary tissue samples demonstrated lectin staining for both SA receptors with α2,6-galactose and α 2,3-galactose linkages.

**Conclusions and Relevance:** The presence of SA receptors that can bind HPAI A H5N1 in human mammary tissue indicates that human milk could be infected during severe avian influenza infection. Pandemic preparedness must prioritize mechanisms to protect the safety of human milk.

## Introduction

Sialic acids (SA) are highly conserved monosaccharides, found across vertebrate species, which serve as attachment sites for multiple viruses including influenza, and are implicated in zoonotic spread of infection from animals to humans.^1^

Highly pathogenic avian influenza (HPAI) A H5N1 (referred to as H5N1 throughout) emerged in 1996 in wild birds and, since 2020, has evolved rapidly to infect a wide range of mammalian species.^2^ In March 2024, the H5N1 clade 2.3.4.4b was identified in dairy cattle in Texas.^3,4^ H5N1 has now been detected in dairy cattle in 17 states impacting more than 1000 herds.^5^ Greater than 70% of California dairy herds were infected between August-December 2024.^5^

Evaluation of infected cattle identified SA receptors in the bovine mammary gland capable of binding both human and avian influenza viruses.^6^ Further analysis demonstrated tropism of the virus for the bovine udder with the highest concentrations of viral RNA found in the udder and in expressed milk.^4,7^ Significantly, raw cow’s milk feeding has been reported as a mechanism for mammal to mammal transmission of H5N1 in a number of species.^3,8^

H5N1 infections in livestock increase interactions between humans and infected animals and raise the risk for spillover. In 2024, 66 confirmed human cases and seven probable cases are reported by the Centers for Disease Control and Prevention (https://www.cdc.gov/bird-flu/situation-summary/index.htmlaccessed01/03/25). Should sustained human to human transmission of H5N1occur, it is imperative to understand the potential for infection of the human mammary gland, as this would have implications for infant feeding.

The objective of this study was to evaluate human mammary tissue for the presence of SA receptors with α2,6- and α2,3-galactose linkages.

## Methods

### Human Subjects Protection

Use of residual deidentified tissue was approved by the University of California, San Diego Health Institutional Review Board

### Tissue Samples

Mammary tissue was selected from residual tissue blocks originally obtained from women of reproductive age, 18-44 years during breast surgery. Lung tissue was obtained in a similar manner and was used as a reference for mammary tissue, as lectin staining for SA receptors is expected to be positive in lung tissue.^1^ All tissue was classified as normal with no evidence of malignancy.

### Lectin Staining

Tissue sections were cut from blocks of formalin-fixed paraffin-embedded breast and lung.Five-micron tissue sections were stained with lectins including biotinylated Maackia Amurensis Lectin 1 (MAL1) and Lectin 2 (MAL2) both of which detect SA in the α 2,3 positions (avian) and Sambucal Nigra Lectin (SNA), which binds preferentially to SA attached to terminal galactose in α-2,6 configuration (human) (Vector Laboratories; Newark, CA).

Slides were stained on a Ventana Discovery Ultra (Ventana Medical Systems, Tucson, AZ). Antigen retrieval was performed using CC1 (Tris-EDTA; pH 8.6) for 40 minutes at 95°C followed by peroxide-blocking using Discovery inhibitor (Ventana). Prior to adding lectins, methods were applied to minimize non-specific staining. Endogenous biotin was blocked using the ReadyProbe Streptavidin/Biotin Blocking system (Invitrogen). Streptavidin was incubated with the tissue to bind endogenous biotin. Following rinsing, remaining biotin sites were blocked by adding biotin for 16 minutes at 22° C. Non-specific protein-binding was blocked using Carbo-free blocking solution (Vector) for one hour.

Bound lectins were detected with SA-HRP (Abcam; 1:250), which was visualized using 3-3’ diaminobenzidine as a chromagen and hematoxylin as a counterstain. Evaluation of staining was performed by two pathologists (HR and SG).

## Results

All breast tissue samples (N=4) demonstrated similar staining across samples and across the three stains. Representative example is shown in Figure 1. Overall, there is patchy luminal ductular epithelium staining (non-nuclear, suggestive of membranous staining). There is scattered basal oriented staining, suggestive of myoepithelial staining. There is staining of luminal secretions. There appears to be more overall ductular epithelium staining for SNA compared with MAL1 and MAL2. There is no appreciable difference in amount of staining between MAL1 and MAL2. SNA, MAL 1, and MAL 2 stained alveolar epithelium in lung tissue as expected, indicating methods used for tissue staining were appropriate.

**Figure 1:**
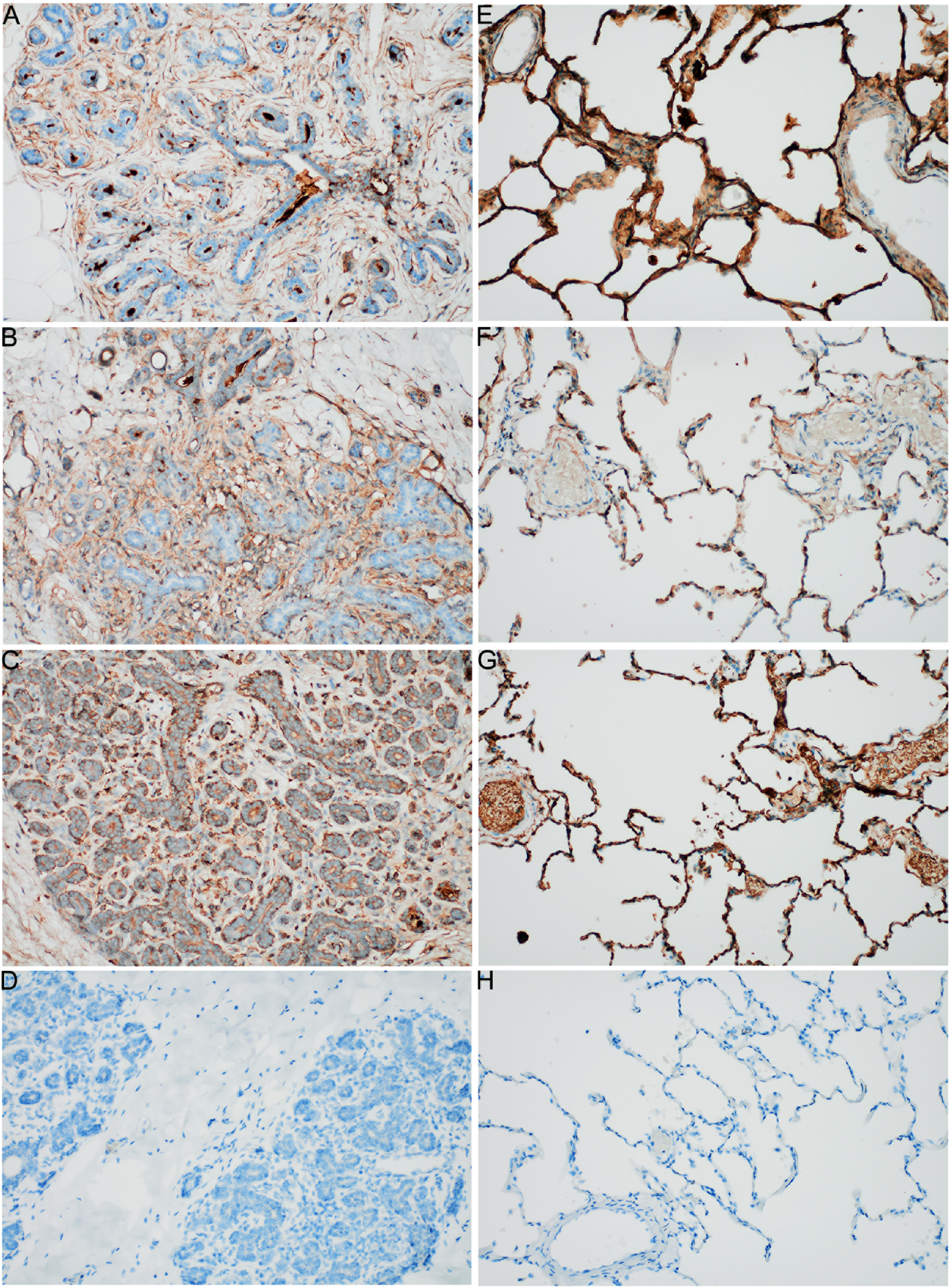
MAL 1, MAL 2, and SNA staining of human mammary and lung tissue. The lectins were labeled with biotin and detected using streptavidin-conjugated horseradish peroxidase. Staining is present within the terminal duct lobular unit in mammary tissue for MAL 1 (A, 200X), MAL 2 (B, 200X), and SNA (C, 200X). Mammary tissue without lectin staining (D, 200X). Staining is present in alveolar epithelium in lung tissue for MAL 1 (E, 200X), MAL 2 (F, 200X), and SNA (G, 200X). Lung tissue without lectin staining (H, 200X).

## Discussion

The H5N1 epizootic is established in the US. The virus is increasingly adapted to mammalian species and outbreaks in dairy cattle have identified milk as a vehicle for viral transmission. Human infants are immunologically immature, at risk for severe viral infections, and reliant on milk feedings. It is therefore imperative to understand the potential for the virus to infect human mammary tissue.

The tropism of influenza viruses depends on the tissue distribution of SA receptors. Previous studies demonstrated SA receptors in humans with α2,6-galactose linkages throughout the respiratory tract and in ileal epithelium, whereas, α 2,3-galactose linkages have been demonstrated in ciliated bronchioles and alveoli, in colon epithelium, in endothelial cells of blood vessels, inflammatory cells, and the conjunctiva.^1,9^ The distribution of SA receptors in human mammary tissue was not reported before the current H5N1 outbreak in dairy cattle.

SA receptors that bind avian influenza were found in udders of dairy cattle, which are analogous, but not identical, to human mammary glands.^3,10^ H5N1 transmission between mammals through milk feeding has been documented. In Argentina, a 95% mortality rate due to H5N1 infection was seen in exclusively nursing elephant seal pups in 2023, with milk identified as a potential vehicle for transmission.^11^ Fatal infections with H5N1 through feeding of infected, raw cow’s milk to cats and mice has also been reported.^3,8^ Experimental infections with avian influenza in ferrets and mice has also resulted in virus identified in mammary tissue and transmission via milk feeding.^12-14^

The current study demonstrated SA receptors with both α2,6- and α 2,3-galactose linkages in human mammary tissue. Another group of investigators recently demonstrated similar SA distribution in the human mammary gland and noted that mammary alveolar epithelium primarily express avian like SA receptors.^15^ Together, these findings indicates that the human mammary gland has the potential to bind avian influenza strains including H5N1 and therefore human milk could be infected.

In humans, seasonal influenza is primarily an upper respiratory disease and viremia, although possible is rare.^16,17^ In contrast, human infections with H5N1 often infect the lower respiratory tract, resulting in more severe disease and are more often associated with viremia and extra-pulmonary manifestations.^16^ Pregnant and nursing women are considered high-risk for severe influenza, including avian influenza.^18^ With increasing mammalian infections, it is critical to understand if human milk can act as a vehicle for transmission of H5N1. Pediatric guidelines do not restrict breast feeding during seasonal influenza infection, but there are no specific recommendations for those infected with H5N1 or other avian strains.^19^ Research examining human milk and emerging influenza viruses, such as was performed during the SARS-CoV-2 pandemic should be prioritized.^20^

### Limitations

The study is limited because mammary tissue samples were obtained from non-lactating women. Cell receptor expression and or binding may change during lactation. importantly, the presence of SA receptors capable of binding avian influenza in mammary tissue does not prove that H5N1 infection of human mammary tissue will occur. Understanding if and under what circumstances infection of milk occurs and whether human milk can transmit virus via oral ingestion should be research priorities to protect infant health prior to the emergence of a pandemic.

In summary, we demonstrated the presence of SA receptors with α2,6-galactose linkages (human) and α 2,3 linkages (avian) in all specimens of mammary tissue evaluated. The presence of receptors is an indication that human mammary tissue is vulnerable to avian influenza infection which could potentially result in virus-contaminated milk. Pandemic preparedness must address needs of all populations, including pregnant women and infants, and include support for the safety of human milk.

## Data Availability

All data produced in the present work are contained in the manuscript

## Acknowledgements

Dr. Byington has intellectual property in and receives royalties from BioFire (biomerieux) through the University of Utah. Dr. Byington is a member of the board of directors of Becton Dickinson.

